# Allele-Specific MicroRNA-Binding Variants at Colorectal Cancer Risk Loci in a Hispanic/Latino Population: An Integrative GWAS, All of Us, and Epigenomic Analysis

**DOI:** 10.64898/2026.09.16.26363169

**Authors:** Sheikh Shafin Ahmad, Md Zahirul Islam Khan, Sourav Roy

**Affiliations:** Department of Environmental Science and Engineering, University of Texas at El Paso, El Paso, TX 79968, USA; The Border Biomedical Research Center, University of Texas at El Paso, El Paso, TX 79968, USA; Department of Biological Sciences, University of Texas at El Paso, El Paso, TX 79968, USA

**Keywords:** colorectal cancer, miRNA, SNP, microRNA binding, Hispanic health disparities, All of Us Research Program

## Abstract

Colorectal cancer (CRC) disproportionately affects Hispanic individuals, who remain underrepresented in genomic discovery and lack ancestry-matched functional resources. Many CRC risk variants are non-coding, and those in 3′ untranslated regions (3′ UTRs) can alter microRNA (miRNA) binding sites and reprogram post-transcriptional gene regulation. Here, we identify allele-specific miRNA-binding variants at CRC risk loci within the Hispanic population using a discovery pipeline anchored to Hispanic-relevant resources. CRC-associated 3′ UTR variants were compiled from the GWAS Catalog, further verified in Hispanic/Latino individuals using the All of Us Research Program cohort (n = 453), expanded into proxy sets using Admixed American-specific linkage disequilibrium, annotated against Ensembl transcripts, and evaluated with TargetScan and RNAhybrid. Screening of 38 CRC-associated 3′ UTR SNPs identified 19 variants that alter miRNA binding across nine genes, with each gene represented by a variant confirmed in the Hispanic cohort. Three oncogenic loci, *DCBLD2* (lead SNP rs11552978), *GREM1* (lead SNP rs10318), and *SMC1B* (rs3747239, prioritized as an AMR-specific proxy in near-complete LD with confirmed lead rs6007600), met all prioritization criteria. In each case, the risk allele disrupts a strong miRNA-binding site, while the gene shows increased tumor expression, weak correlation between expression and methylation, and an absence of local active transcriptional chromatin marks (cCREs/H3K27Ac). These results suggest that the variants promote gene overexpression through loss of miRNA-mediated repression rather than changes in promoter methylation. Overall, the study identifies non-coding CRC susceptibility variants in the Hispanic population and highlights *DCBLD2, GREM1*, and *SMC1B* as strong candidates for functional validation.

## 1. Introduction

Colorectal cancer (CRC) is the third most frequently diagnosed malignancy worldwide and the second leading cause of cancer death, with an estimated 1.93 million new cases and 904,000 deaths in 2022 (Siegel et al. 2023; Wu et al. 2025). Hispanic and Latino individuals in the United States are frequently diagnosed at later stages, show a rising incidence of early-onset cases, and experience poorer stage-adjusted survival. These disparities are not uniform across the Hispanic population as incidence varies substantially by country of origin, reflecting the diversity often overlooked in the single “Hispanic” category in U.S. cancer registries (Stern et al. 2015). These disparities come from a complex mix of socioeconomic barriers, environmental factors, and ancestral genetic factors that remain poorly resolved at the molecular level. The goal of this study is to identify allele-specific miRNA-binding variants at CRC risk loci in the Hispanic population using Hispanic-specific genetic datasets throughout the discovery pipeline, rather than by comparison with other populations.

At the molecular level, CRC has long been understood through sequential mutations in key genes, including *APC, KRAS*, and *TP53*, which drive the adenoma-to-carcinoma sequence (Fearon and Vogelstein 1990). This framework has been indispensable for understanding somatic, coding-sequence drivers, but it describes little about inherited, non-coding susceptibility variants, which act through fundamentally different regulatory mechanisms and are the focus of the present study.

Genome-Wide Association Studies (GWAS) have served as a powerful approach for identifying genomic loci associated with colorectal cancer susceptibility across broad populations (Tenesa and Dunlop 2009). However, since historical GWAS have relied almost entirely on cohorts of European descent (Popejoy and Fullerton 2016), their diagnostic power leaves a massive blind spot when applied to Admixed American (AMR) populations. Because genetic backgrounds differ across populations, risk prediction tools developed from European-based studies often perform less accurately in AMR populations. For example, polygenic risk scores often show reduced accuracy when applied to populations with genetic backgrounds that differ from the populations in which they were developed, limiting their clinical utility and potentially contributing to existing health disparities (Martin et al. 2019; Ding et al. 2023).

Differences in linkage disequilibrium (LD) patterns, which are shaped by population history, genetic variation, and admixture, further complicate the interpretation of genetic risk variants across populations (Auton et al. 2015). A variant associated with disease risk in European populations may therefore not accurately represent the same biological signal in Hispanic individuals. Multi-ethnic GWAS efforts, such as the Population Architecture using Genomics and Epidemiology (PAGE) study, have demonstrated that incorporating diverse populations improves the discovery of disease-associated loci and helps more accurately identify the variants most likely to drive disease risk (Wojcik et al. 2019). However, functional genomic resources, including expression quantitative trait locus (eQTL) datasets, remain largely based on European ancestry and non-cancer tissues, leaving a major gap in ancestry-matched functional data for Hispanic CRC (Ardlie et al. 2015).

The *All of Us* Research Program assembles a massive genomic library to address this limitation where over half the data comes from non-European backgrounds (Bick et al. 2024). By evaluating GWAS-identified risk loci within the ancestral background of the *All of Us* Hispanic cohort, our integrative framework facilitates the identification of functional, post-transcriptional regulatory variants that may be overlooked in European-centric models.

Another challenge is that a huge portion of the cancer risk variants found in GWAS lies in non-coding sequence, where their functional consequences are not obvious. Among these, variants in the 3′ untranslated regions (3′ UTRs) of protein-coding genes are increasingly recognized as important post-transcriptional regulators. MicroRNAs (miRNAs) are short, roughly 22-nucleotide, non-coding RNAs that bind complementary seed sequences in target 3′ UTRs and trigger mRNA destabilization or translational repression (Bartel 2009). A single base substitution due to a reported SNP at such a site can either remove a miRNA-mRNA binding site, causing the target to be over-expressed, or create a new site, imposing gene downregulation. These are commonly known as mirSNP. This mechanism has already been demonstrated in CRC, where SNPs rs61764370 and rs712 in the *KRAS* 3′ UTR were shown to increase CRC risk by disrupting binding of let-7 impacting KRAS transcription (Dai et al. 2015).

The functional impact of mirSNP also depends on the surrounding epigenetic state. The local chromatin state, including DNA methylation at CpG islands and histone modifications such as H3K27 acetylation (H3K27Ac), manages whether a locus is transcriptionally active and whether its 3′ UTR is even available to the miRNA machinery (Dunham et al. 2012; Roadmap Epigenomics Consortium et al. 2015). A loss-of-binding variant only matters if the host gene is actively transcribed, and a gain-of-binding variant matters when the gene is being silenced. Therefore, interpreting mirSNPs requires reading genotype and epigenome together, not just genotype alone.

Our work builds on a previous study that successfully linked non-coding genetic risks in breast cancer to disrupted microRNA bindings (Jacinta-Fernandes et al. 2020). We expand on that strategy here by extending it to an underrepresented population by explicitly accounting for AMR genomic architecture. We assembled an integrative, population-specific pipeline that combines GWAS Catalog and All of Us variant discovery with AMR-specific LD mapping, Ensembl 3′ UTR annotation, dual thermodynamic miRNA-binding modelling, and epigenomic and survival context. Applying this pipeline to Hispanic/AMR CRC loci, we narrow a large candidate pool down to a small set of high-confidence functional variants and focus on three oncogenes, *DCBLD2, GREM1*, and *SMC1B*, that satisfy several independent criteria at once. They showed the expected increase in tumor expression, showed a low correlation with DNA methylation, and lacked confounding local transcriptional promoter/enhancer marks. These represent the strongest mechanistic leads for downstream functional work.

## 2. Materials and Methods

Figure 1 gives an overview of the four-stage pipeline: variant discovery, annotation and mapping, in-silico binding modelling, and validation and prioritization.

**Fig. 1.**
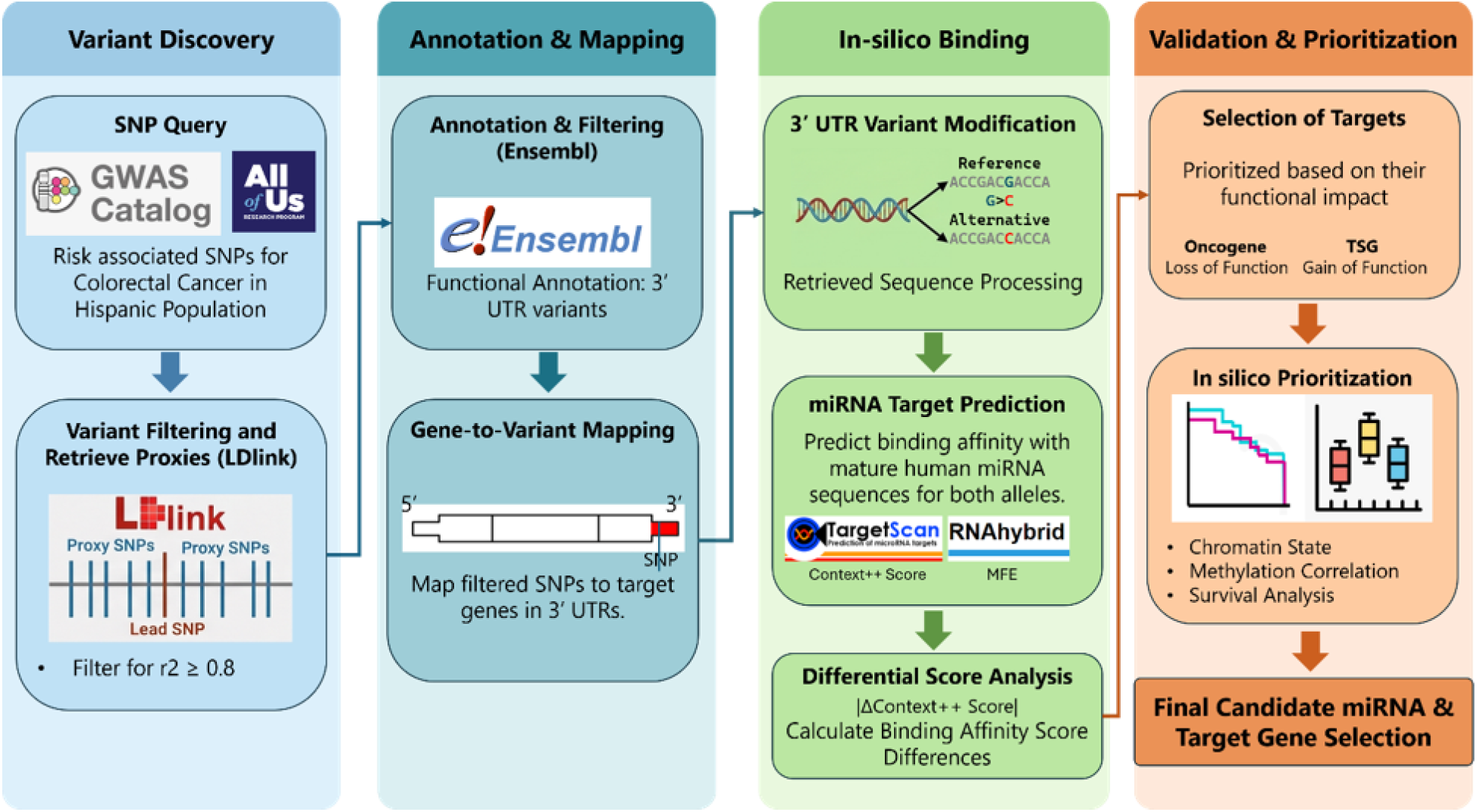
Overview of the discovery pipeline used to identify allele-specific microRNA-binding variants at CRC risk loci in the Hispanic population. CRC-associated SNPs were compiled from the GWAS Catalog and checked for presence in a Hispanic/Latino cohort from the All of Us Research Program, then expanded into LD proxies within the AMR reference population. Proxy variants were annotated and mapped to their nearest genes, retaining only those located in the 3′ untranslated region (3′ UTR) of protein-coding transcripts, since this is where microRNAs bind. The reference and risk (alternative) alleles of each variant were then compared using two independent microRNA-target-prediction tools to identify variants that gain or lose a microRNA binding site. Finally, candidate variants were ranked using tumor chromatin state, DNA methylation, and patient survival data to identify the most biologically plausible risk variants for follow-up

### 2.1. Variant Discovery and Population-Level Confirmation

To build our initial panel of genetic variants, we queried the NHGRI-EBI GWAS Catalog using the broad search term **“Colorectal cancer”** to compile an inventory of established, risk-associated seed SNPs (Buniello et al. 2019). This database served as the initial source of candidate variants evaluated in this study, resulting in 38 unique SNPs that are in the 3′ UTR, representing 45 SNP–trait associations (Supplementary Table S1). Because many GWAS discoveries are derived primarily from European-ancestry populations and may not be present or common in Hispanic/Latino populations, each candidate SNP was independently evaluated for its presence in the Hispanic/Latino population using the *All of Us* Research Program (Controlled Tier Dataset v8; Curated Data Repository [CDR] accessed February 2026) (Bick et al. 2024). A Hispanic/Latino CRC cohort was identified within this Researcher Workbench using three selection criteria: (1) an electronic health record (EHR) clinical diagnosis code for “Malignant Neoplasm of Colon and/or Rectum” (*n* = 4,110), (2) self-reported ethnicity for “Hispanic or Latino” (*n* = 112,751), and (3) available short-read whole-genome sequencing (srWGS) data (*n* = 414,830) (Table 1). This intersection identified a final population confirmation cohort of 453 individuals (Supplementary Table S2). Age at the time of data curation spanned three primary brackets: 18–44 years (*n* = 36, 8%), 45–64 years (*n* = 213, 47%), and >64 years (*n* = 204, 45%). All reported aggregate counts strictly adhere to the *All of Us* Data and Statistics Dissemination Policy.

**Table 1.** Definition and Composition of the Hispanic/Latino Colorectal Cancer Confirmation Cohort (*n* = 453)

| Parameter/Criterion | Inclusion/Data Specification | Participants (n/%) |
| --- | --- | --- |
| **Phenotype** | Clinical condition concept:<br>*Malignant neoplasm of colon and/or rectum* | 4,110 total cases |
| **Ethnicity** | *Hispanic or Latino* | 112,751 total participants |
| **Genomic Data Modality** | Short-Read Whole Genome Sequencing (srWGS) | 414,830 total sequenced in dataset |
| **Final Evaluated Cohort** | Intersection of Diagnosis ∩ Hispanic Ethnicity ∩ srWGS | $n = 453$ |
| **Age at Current CDR (years)** |  |  |
| 18–44 |  | 36 (8%) |
| 45–64 |  | 213 (47%) |
| >64 |  | 204 (45%) |
| **Data Release** | All of Us Research Program, Controlled Tier Dataset v8 |  |

In this computational pipeline, the *All of Us* cohort served strictly as an independent population-level genomic filter to confirm variant carriage in sequenced Hispanic CRC patients, rather than for clinical staging or epidemiological subgroup analysis. SNPs confirmed in this Hispanic cohort were cross-referenced with the GWAS pool, yielding a refined set of high-priority 3′ UTR risk loci for downstream linkage disequilibrium mapping, thermodynamic modeling, and functional prioritization.

### 2.2. Linkage-Disequilibrium Analysis and Proxy Extraction

Since GWAS-identified lead SNPs often represent genetic markers linked to the true functional variant rather than the causal variant itself, we performed LD-based proxy analysis using the LDlink suite (LDproxy and LDpair) (Machiela and Chanock 2015). For each lead SNP, we identified correlated variants located within a ±500 kb genomic region. LD calculations were performed using the AMR reference population from the 1000 Genomes Project (Auton et al. 2015) to better capture population-specific genetic relationships and haplotype patterns in Hispanic/Latino populations. This approach is supported by previous studies demonstrating that LD mapping using diverse and admixed populations can improve the identification and fine-mapping of potential causal variants compared with analyses based primarily on European-ancestry reference panels (Wojcik et al. 2019). Following those studies, we too applied a stringent compound linkage disequilibrium threshold of r^2^ ≥ 0.8 and D′ = 1.0. While r^2^ measures correlation in allele frequencies, requiring D′ = 1.0 specifically ensures complete historical co-inheritance without evidence of historical recombination events between the lead GWAS marker and the candidate functional variant within the Admixed American (AMR) haplotype background, minimizing false-positive proxy selection. These variants showing strong linkage with the lead SNPs (r^2^ ≥ 0.8 and D′ = 1) were selected as LD proxies and included in subsequent functional annotation analyses (Supplementary Table S3).

### 2.3. Structural Annotation and Gene Mapping

The expanded SNP pool was annotated using Ensembl BioMart (GRCh38) tool and filtered to variants mapping exclusively to 3′ UTR coordinates of protein-coding transcripts. Each retained variant was assigned to its host gene, strand and reference/alternative alleles. Its putative CRC role (upregulated or downregulated target) was also recorded from previous literature (Supplementary Table S4).

### 2.4. In-Silico miRNA–mRNA Binding and Thermodynamic Modelling

For each candidate 3′ UTR variant, the full 3′ UTR sequence of the host transcript was retrieved from Ensembl. At the variant’s genomic coordinate, the reference nucleotide was substituted with the alternative allele to generate matched reference and alternative allele sequences. Mature human miRNA sequences were obtained from miRBase (v22) (Kozomara and Griffiths-Jones 2014). Following an established dual-modelling strategy by Jacinta-Fernandes et al. (Jacinta-Fernandes et al. 2020), allele-specific binding was evaluated with two complementary tools:

**TargetScan (v8.0)** computed the context++ score, a quantitative estimate of a site’s repressive effect that integrates local AU content, site position, and pairing features (Agarwal et al. 2015). More negative scores indicate stronger predicted repression.

**RNAhybrid (v2.1.2)** computed the minimum free energy (MFE, kcal/mol) of the predicted miRNA–mRNA duplex, giving a thermodynamic measure of hybridization stability (Rehmsmeier et al. 2004).

Allele-specific effects were summarized as the differential context++ score, ΔContext++ = Context++(Alt) − Context++(Ref). A positive value indicates loss-of-binding (the reference site is repressed but the alternative allele escapes the miRNA), whereas a negative value indicates gain-of-binding (the alternative allele creates a novel, stable site). Per-site binding details for all prioritized genes are provided in Supplementary Table S5.

### 2.5. Epigenomic Context, Expression, Methylation, and Survival

To evaluate the biological plausibility of our prioritized variants, we examined tissue-level chromatin structures, gene expression patterns, and DNA methylation profiles from publicly available datasets. It should be noted that available public multi-omic resources are predominantly European-centric and not ancestry-matched to Hispanic cohorts. We leverage these tools strictly to establish the general carcinogenic potential and functional landscape of our candidate genes, providing crucial biochemical validation that supports our population-specific claims:

#### Chromatin state

We used the UCSC Genome Browser (GRCh38/hg38) to inspect active histone marks focusing on H3K27Ac enrichment, alongside candidate cis-regulatory element (cCRE) annotations across primary colon tissues and colorectal cancer (CRC) cell lines. By leveraging public ENCODE and Roadmap Epigenomics tracks (Dunham et al. 2012; Roadmap Epigenomics Consortium et al. 2015), we map these features to assess local chromatin accessibility and rule out confounding promoter or enhancer activity.

#### Expression, methylation and survival

Tumor-versus-normal expression and expression–methylation correlation for prioritized genes were obtained from OncoDB (Tang et al. 2022), an integrated database of cancer gene expression and DNA methylation derived from TCGA and related cohorts. A negligible expression–methylation correlation for an over-expressed oncogene was interpreted as evidence favoring a post-transcriptional (miRNA-mediated) mechanism rather than epigenetic promoter control. Expression-stratified overall survival was assessed independently using the KM Plotter pan-cancer resource (Győrffy 2024), which generates Kaplan–Meier curves and log-rank statistics from microarray and RNA-seq cohorts stratified by median target-gene expression.

Statistical significance for differential expression was assessed using Student’s t-tests, expression–methylation associations were evaluated using Pearson correlation coefficients, and survival differences were tested via log-rank tests. Because this multi-stage bioinformatic pipeline was designed as an exploratory, hypothesis-generating prioritization framework integrating orthogonal genomic layers rather than simultaneous parallel hypothesis testing across thousands of unselected features, unadjusted nominal p-values are reported alongside exact effect sizes and 95% confidence intervals. Nonetheless, candidate gene differential expression tests remained significant after applying False Discovery Rate (FDR) Benjamini–Hochberg correction across the evaluated candidate gene set (q < 0.001 for DCBLD2, GREM1, and SMC1B).

### 2.6. Variant Prioritization Criteria

To narrow the initial pool of candidate 3′ UTR variants down to high-confidence functional drivers, variants were required to satisfy five hierarchical inclusion criteria:

- **Allele-Specific Binding Disruption:** Complete or substantial loss of predicted miRNA binding (|ΔContext++ score| ≥ 0.5 or loss of 7mer/8mer seed pairing) or novel stable site creation evaluated by TargetScan and RNAhybrid.
- **Population Relevance:** Direct confirmation in the All of Us Hispanic/Latino cohort (n = 453) or near-complete LD (r^2^ ≥ 0.8, D′ = 1.0) with a confirmed lead SNP in the 1000G AMR panel.
- **Concordant Tumor Overexpression:** Statistically significant upregulation of the host gene in colorectal adenocarcinoma (COAD) tissues compared to matched normal tissues in OncoDB (p < 0.05).
- **Independence from Promoter Methylation:** Negligible or weak correlation between mRNA expression and promoter DNA methylation (|R| < 0.3), indicating that upregulation is driven by post-transcriptional evasion rather than promoter hypomethylation.
- **Clinical Outcome Association:** Significant association between elevated host gene expression and reduced patient overall survival in Kaplan–Meier survival analysis (log-rank p < 0.05).

## 3. Results

### 3.1. Validation of the computational pipeline using experimentally verified miRSNPs

Before investigating CRC-associated variants, we first evaluated the accuracy of our computational pipeline using previously validated miRNA-binding SNPs (miR-SNPs). This validation was performed to determine whether the combined TargetScan and RNAhybrid workflow could reliably identify allele-specific changes in miRNA binding that had been experimentally confirmed. Successful reproduction of known functional variants would support the use of this strategy for discovering novel CRC-associated miR-SNPs. Six experimentally validated miR-SNPs associated with different human diseases were analyzed (Table 2), including examples of both loss and gain of miRNA binding. For each variant, the reference and alternative alleles were evaluated using TargetScan to predict regulatory effects (context++ score) and RNAhybrid to estimate the thermodynamic stability of miRNA–mRNA interactions through minimum free energy (MFE).

**Table 2.** Previously functionally validated SNPs affecting miRNA binding sites, recovered by the TargetScan/RNAhybrid pipeline.

| SNP ID | Gene | miRNA | Alleles | Ref<br>Context<br>++ | Ref<br>MFE | Alt<br>Context<br>++ | Alt<br>MFE | Associated<br>disease | Ref |
| --- | --- | --- | --- | --- | --- | --- | --- | --- | --- |
| **rs12720208** | *FGF20* | hsa-miR-433-3p | G/A | -0.941 | — | — | — | Parkinson's disease | (Wang et al. 2008) |
| **rs5186** | *AGTR1* | hsa-miR-155 | A/C | -0.894 | — | — | — | Hypertension | (Haas et al. 2012) |
| **rs10876135** | *TFCP2* | hsa-miR-197-5p | A/G | -0.825 | — | — | — | Alzheimer's disease | (Roy and Mallick 2017) |
| **rs3742943** | *JAG2* | hsa-miR-1224-3p | C/T | -0.679 | — | — | — | Neurodegeneration | (Saba et al. 2014) |
| **rs9291296** | *GABRA4* | hsa-miR-26a-5p | A/G | — | — | -1.767 | — | Autism | (Saba et al. 2014) |
| **rs11540855** | *ABHD8* | hsa-miR-4707-3p | A/G | — | — | -0.992 | -27.5 | Breast cancer | (Li et al. 2017b) |

*Context++* scores are unitless (*TargetScan*); MFE in kcal/mol (*RNAhybrid*). Blank cells(“—”) indicate *TargetScan* predicted no binding site for that allele (*Context++* score not applicable); and for MFE it means the value did not meet *RNAhybrid*'s stability/significance cutoff.

**Table 3.** Hispanic/Latino All of Us cohort confirmation status for all 19 candidate variants across the nine prioritized genes. Every gene has at least one directly confirmed lead variant (bold); secondary and proxy variants within a gene are resolved via AMR-panel LD (Section 2.2)

| Gene | Variant(s) screened | Confirmed in All of Us Hispanic cohort |
| --- | --- | --- |
| **TTC22** | rs12144319 | Yes |
| **C2orf72** | rs73089558 | Yes |
| **VIPR1** | rs8913, rs895, rs897 | Yes (rs8913); rs895/rs897 via AMR LD |
| **DCBLD2** | rs11552978 | Yes |
| **NT5E** | rs6913634 | Yes |
| **ITPR2** | rs2570, rs8311 | Yes (rs2570); rs8311 via AMR LD |
| **SMAD9** | rs511674, rs514501, rs517297, rs518086, rs671860, rs683296, rs17054567 | Yes (rs511674); remaining 5 via AMR LD |
| **GREM1** | rs10318 | Yes |
| **SMC1B** | rs6007600, rs3747239 | Yes (rs6007600); rs3747239 via AMR LD |

The computational pipeline successfully reproduced all six previously validated miRNA-binding events, demonstrating its ability to accurately identify allele-specific changes in miRNA binding. For example, the Parkinson’s disease-associated variant *FGF20* rs12720208 showed a strong predicted binding site for hsa-miR-433-3p in the reference allele (context++ = −0.941), whereas the alternative allele abolished this interaction, consistent with the experimentally validated loss of miRNA-mediated regulation (Wang et al. 2008). Conversely, the breast cancer-associated variant *ABHD8* rs11540855 created a novel binding site for hsa-miR-4707-3p in the alternative allele, with a strong context++ score (−0.992) and a stable minimum free energy of −27.5 kcal/mol, reflecting a previously reported gain-of-binding event (Li et al. 2017b). Similar agreement was observed for *AGTR1, TFCP2, GABRA4*, and *JAG2* (Table 2), indicating that the workflow consistently identified both disrupted and newly created miRNA-binding sites. Collectively, these findings validate the robustness of the combined TargetScan/RNAhybrid approach and support its application for the systematic identification of functional CRC-associated miRNA-binding variants in the Hispanic population.

### 3.2. Genome-wide screening identifies 19 candidate miRNA-binding variants across nine CRC susceptibility genes in the Hispanic population

Following validation of the computational pipeline, we applied the same workflow to identify functional miRNA-binding variants associated with CRC susceptibility in the Hispanic population. A total of 38 unique CRC-associated 3′ UTR SNPs, representing 45 SNP–trait associations, were initially retrieved from the GWAS Catalog (Supplementary Table S1). To determine their relevance in the target population, each variant was examined in the All of Us Hispanic/Latino cohort, where 22 variants were directly detected among 453 individuals (Supplementary Table S2). Rather than serving as a strict filtering step, confirmation in the Hispanic cohort provided population-specific support for variants previously identified through GWAS.

To capture potential functional variants that may not represent the original GWAS lead SNPs, the confirmed loci were expanded using AMR-specific LD analysis. Subsequent structural annotation identified variants located within the 3′ UTRs of protein-coding genes, the primary binding sites for miRNAs. Allele-specific binding was then evaluated using both TargetScan and RNAhybrid, allowing comparison of the predicted regulatory effects between reference and alternative alleles (Supplementary Tables S3–S5).

This integrative screening strategy identified 19 candidate miRNA-binding variants across nine genes, including six genes previously reported to be upregulated in CRC (TTC22, DCBLD2, NT5E, GREM1, SMC1B, and C2orf72) and three genes with reported tumor-suppressive functions (VIPR1, ITPR2, and SMAD9) (Supplementary Tables S4–S6). Importantly, each of these nine candidate genes was represented at the locus level by at least one GWAS lead variant directly confirmed in the Hispanic/Latino All of Us cohort. For eight loci, the candidate variants were directly verified, whereas for SMC1B, the candidate functional variant (rs3747239) was resolved through near-complete linkage disequilibrium (r^2^ = 0.994, D′ = 1) with the directly confirmed lead marker rs6007600 within the AMR panel. These findings substantially reduced the initial set of CRC-associated variants to a focused panel of biologically relevant candidate miRNA-binding variants for downstream functional prioritization.

### 3.3. Multi-level functional evidence prioritizes three high-confidence CRC loci

From the 19 candidate miRNA-binding variants identified across nine CRC-associated genes, only three genes, *DCBLD2, GREM1*, and *SMC1B*, consistently demonstrated evidence supporting a functional role in CRC (Table 4). The prioritized variants showed complete or near-complete loss of predicted miRNA binding, and all three genes were significantly overexpressed in CRC tumors compared with their normal tissues. High expression of each gene was also associated with poorer overall survival, supporting their potential clinical relevance. Furthermore, *DCBLD2* and *GREM1* exhibited weak or negligible correlations between gene expression and promoter methylation, suggesting that loss of miRNA-mediated regulation, rather than promoter methylation, is the predominant mechanism underlying their overexpression. Although *SMC1B* also showed loss of miRNA binding and increased tumor expression, its moderate inverse correlation with methylation indicates that both epigenetic and post-transcriptional mechanisms may contribute to its regulation. In contrast, the remaining six candidate genes lacked consistent support across these analyses and were therefore retained as computational candidates for future investigation (Supplementary Table S6).

**Table 4.** Allele-specific miRNA binding for the three prioritized oncogenes.

| GWAS Lead SNP | Candidate SNP | Gene | Ref\Alt | Predicted miRNA target | Ref Con++ | Ref MFE | Alt Con++ | Alt MFE |
| --- | --- | --- | --- | --- | --- | --- | --- | --- |
| **rs11552978** | **rs11552978** (lead) | *DCBLD2* | G\A | hsa-miR-211-3p | -0.963 | -21.6 kcal/mol | — | — |
| **rs10318** | **rs10318** (lead) | *GREM1* | C\T | hsa-miR-331-5p | -0.999 | — | — | — |
| **rs6007600** | **rs3747239** (proxy; $r^2 = 0.994$, $D' = 1$) | *SMC1B* | A\C | hsa-miR-4452 | -0.952 | — | — | — |

*TargetScan context++ score (Con++) is unitless and “—” is used to show it returned no value for the specific mRNA-miRNA; and for MFE, it indicates value fell outside the thermodynamic stability thresholds (MFE) or minimum statistical significance criteria of the RNAhybrid algorithm.*

#### 3.3.1. The risk allele of rs11552978 abolishes hsa-miR-211-3p binding and is associated with increased DCBLD2 expression

The lead variant rs11552978, located within the 3′ UTR of *DCBLD2*, emerged as the highest-confidence candidate identified in this study. Allele-specific binding analysis showed that the reference G allele formed a strong binding site for hsa-miR-211-3p (context++ = −0.963; MFE = −21.6 kcal/mol), whereas the alternative risk A allele completely abolished this interaction, indicating loss of miRNA-mediated regulation. Consistent with this prediction, *DCBLD2* was significantly overexpressed in colorectal tumor tissues compared with normal colon tissues (p = 7.6 × 10^−6^; Figure 2a). In addition, patients with high *DCBLD2* expression had significantly poorer overall survival than those with low expression (HR 1.75, 95% CI 1.38–2.21; log-rank p = 2.1 × 10^−6^; Figure 2b), supporting its association with an aggressive disease phenotype.

**Fig. 2.**
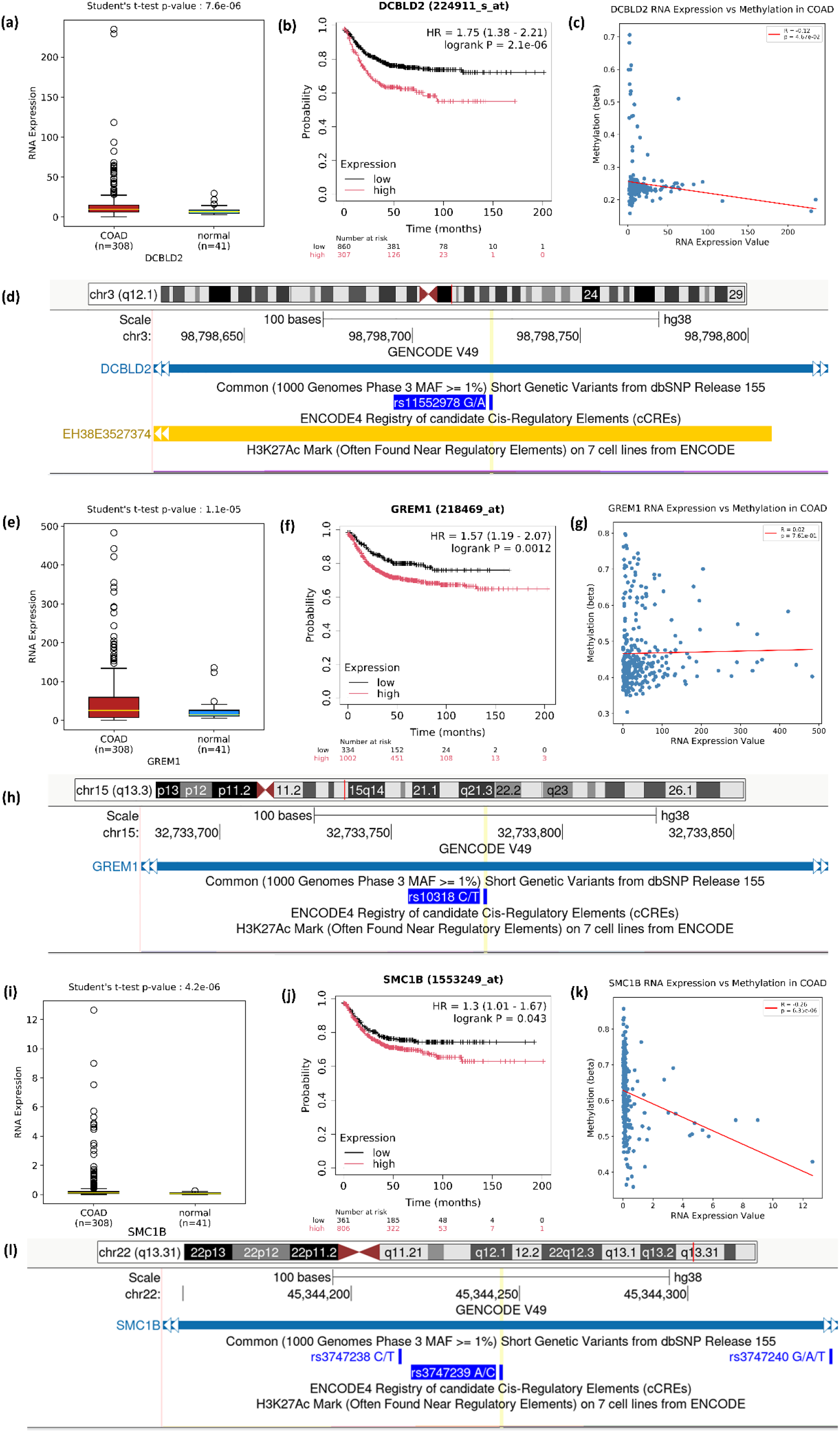
Comprehensive expression, survival, and epigenomic characterization of DCBLD2, GREM1, and SMC1B in colorectal adenocarcinoma. For the DCBLD2 locus, (a) RNA expression is significantly elevated in COAD (n = 308) compared to matched normal tissue (n = 41; Student’s t-test p = 7.6 × 10^−6^), (b) high expression correlates with reduced overall survival (HR = 1.75, 95% CI: 1.38–2.21, log-rank p = 2.1 × 10^−6^), (c) expression shows a weak negative correlation with DNA methylation (R = −0.12, p = 4.67 × 10^−2^), and (d) the UCSC Genome Browser track (GRCh38/hg38) centered on 3′ UTR variant rs11552978 displays tracks for ENCODE4 candidate cis-regulatory elements (cCREs) and the H3K27Ac histone mark, with no active H3K27Ac signals visualized in this region. For the GREM1 locus, (e) RNA expression is increased in tumor tissue (p = 1.1 × 10^−5^), (f) high expression is associated with poorer overall survival (HR = 1.57, 95% CI: 1.19–2.07, log-rank p = 0.0012), (g) no expression–methylation correlation is observed (R = 0.02, p = 7.61 × 10^−1^), and (h) the UCSC Genome Browser track around rs10318 shows no visualized ENCODE4 cCREs or H3K27Ac signals within the genomic window. For the SMC1B locus, (i) RNA expression is significantly higher in COAD (p = 4.2 × 10^−6^), (j) high expression is linked to worse survival outcomes (HR = 1.30, 95% CI: 1.01–1.67, log-rank p = 0.043), (k) expression displays a statistically significant negative correlation with methylation (R = −0.26, p = 6.35 × 10^−6^), and (l) the UCSC Genome Browser track around rs3747239 shows no visualized ENCODE4 cCREs or H3K27Ac signals across the displayed region

Expression–methylation analysis demonstrated only a weak correlation between *DCBLD2* expression and promoter methylation (R = −0.12; Figure 2c), suggesting that promoter methylation is unlikely to be the primary regulator of its increased expression. Examination of variants in perfect linkage disequilibrium (r^2^ = 1.0) showed that these proxies were either synonymous or non-coding variants without predicted functional consequences, further supporting rs11552978 as the most likely functional variant at this locus. Chromatin analysis also showed no candidate cis-regulatory elements (cCREs) and minimal H3K27Ac enrichment surrounding the variant (Figure 2d), indicating that the observed association is unlikely to be explained by local promoter or enhancer activity. Previous studies have shown that miR-211-3p regulates colorectal cancer progression through CHD5 (Cai et al. 2012), while *DCBLD2* has been implicated in epithelial–mesenchymal transition (EMT), invasion, and metastasis across multiple cancers (Xie et al. 2021). Together, these findings identify rs11552978 as a strong candidate miRNA-binding variant that may contribute to CRC susceptibility through allele-specific dysregulation of *DCBLD2*.

#### 3.3.2. The rs10318 risk allele disrupts hsa-miR-331-5p binding and is associated with GREM1 overexpression

The *GREM1* variant rs10318 showed one of the strongest allele-specific effects identified in this study. The reference C allele formed a strong binding site for hsa-miR-331-5p (context++ = −0.999), whereas the alternative risk T allele completely abolished this interaction, indicating loss of miRNA-mediated regulation. Consistent with this prediction, *GREM1* expression was significantly higher in colorectal tumors than in normal colon tissue (p = 1.1 × 10^−5^; Figure 2e). Increased *GREM1* expression was also associated with significantly poorer overall survival in CRC patients (HR 1.57, 95% CI 1.19–2.07; log-rank p = 0.0012; Figure 2f).

Expression–methylation analysis demonstrated a negligible correlation between *GREM1* expression and promoter methylation (R = 0.02; Figure 2g), suggesting that promoter methylation is unlikely to account for its elevated expression. Chromatin analysis also revealed an absence of ENCODE4 candidate cis-regulatory elements (cCREs) and no detectable H3K27Ac enrichment across the variant locus (Figure 2h), indicating that the observed upregulation is unlikely to be driven by local transcriptional enhancer or promoter activity and instead supports a post-transcriptional mechanism. Previous studies have established *GREM1* as a CRC susceptibility gene involved in BMP signaling (Jaeger et al. 2008; Tomlinson et al. 2011), and another functional 3′ UTR variant has been reported to increase CRC risk by disrupting hsa-miR-185-3p binding (Li et al. 2017a). Together, these findings identify rs10318 as an additional candidate miRNA-binding variant that may contribute to colorectal cancer susceptibility through allele-specific upregulation of *GREM1*.

#### 3.3.3. The rs3747239 risk allele alters miRNA binding and is associated with increased SMC1B expression

The candidate variant rs3747239, identified through AMR-specific linkage disequilibrium with the lead GWAS marker rs6007600 (r^2^ = 0.994, D′ = 1), demonstrated a marked allele-specific change in miRNA binding. The reference A allele formed a strong binding site for hsa-miR-4452 (context++ = −0.952), whereas the alternative C allele abolished this interaction and simultaneously generated a predicted binding site for hsa-miR-20a-3p (context++ = −0.91; Supplementary Table S5). Consistent with these predictions, SMC1B expression was significantly higher in colorectal tumor tissues than in normal tissues (p = 4.2 × 10^−6^; Figure 2i), and elevated expression was associated with poorer overall survival (HR 1.30, 95% CI 1.01–1.67; log-rank p = 0.043; Figure 2j).

Unlike *DCBLD2* and *GREM1, SMC1B* showed an inverse correlation between gene expression and promoter methylation (R = −0.26, p = 6.35 × 10^−6^; Figure 2k), suggesting that both epigenetic and post-transcriptional mechanisms may contribute to its regulation. Chromatin analysis showed no visualized ENCODE4 cCREs or H3K27Ac enrichment across the variant locus (Figure 2l), arguing against a local promoter or enhancer role and supporting a post-transcriptional regulatory mechanism. Previous studies have implicated aberrant expression of SMC1B and other cohesin components in chromosomal instability and tumor progression (Losada 2014; Sarogni et al. 2019; Boukaba et al. 2022). Collectively, these findings support rs3747239 as a functional candidate variant that may contribute to CRC susceptibility through altered miRNA-mediated regulation of *SMC1B*.

## 4. Discussion

In this study, we developed an ancestry-anchored functional genomics pipeline to prioritize non-coding CRC susceptibility variants that alter post-transcriptional miRNA regulation in Hispanic populations. By integrating GWAS loci, Hispanic-cohort confirmation via *All of Us*, AMR-specific linkage disequilibrium mapping, dual thermodynamic miRNA-binding predictions, and multi-omic validation, we identified *DCBLD2* (rs11552978), *GREM1* (rs10318), and *SMC1B* (rs3747239) as candidate functional drivers. Unlike traditional association studies that do not elucidate non-coding regulatory mechanisms, our findings provide a mechanistic framework linking inherited 3′ UTR variants to allele-specific miRNA evasion and oncogenic overexpression in an underrepresented population.

Although all three prioritized genes converge on a common mechanism involving disruption of miRNA binding, each represents a distinct biological pathway implicated in colorectal tumorigenesis. *DCBLD2* has been associated with epithelial-mesenchymal transition (EMT), invasion, and metastasis in gastrointestinal and several other cancers (Xie et al. 2021). Our analyses indicate that the risk allele of rs11552978 abolishes hsa-miR-211-3p binding while occurring in a genomic region with minimal evidence of promoter or enhancer activity. Together with the weak correlation between gene expression and promoter methylation, these findings support the hypothesis that altered post-transcriptional regulation, rather than epigenetic silencing, contributes to *DCBLD2* overexpression. Similarly, GREM1, a well-established CRC susceptibility gene involved in BMP signaling (Tomlinson et al. 2011), showed complete loss of hsa-miR-331-5p binding associated with the risk allele. This observation complements previous evidence demonstrating that disruption of another miRNA-binding site (hsa-miR-185-3p) can increase CRC risk through GREM1 dysregulation (Li et al. 2017a), suggesting that multiple independent miRNA-mediated mechanisms may regulate this locus. In contrast, although SMC1B also demonstrated allele-specific disruption of miRNA binding, its stronger association with promoter methylation indicates that both epigenetic and post-transcriptional mechanisms may contribute to its regulation. Given the established role of cohesin dysregulation in chromosomal instability and colorectal tumorigenesis (Losada 2014; Sarogni et al. 2019; Boukaba et al. 2022), altered regulation of SMC1B represents another plausible mechanism underlying inherited CRC susceptibility.

One major strength of this study is the incorporation of ancestry-specific genetic information throughout the discovery process. Most CRC susceptibility loci have been identified in populations of predominantly European ancestry, limiting their direct applicability to admixed populations such as Hispanics (Popejoy and Fullerton 2016; Martin et al. 2019; Ding et al. 2023). Rather than relying exclusively on published GWAS findings, we confirmed the presence of candidate variants in Hispanic participants from the *All of Us* Research Program and performed linkage disequilibrium mapping using the AMR reference panel. This population-specific strategy proved particularly valuable for SMC1B, where we identified the prioritized functional variant (rs3747239) through its near-complete LD with the lead GWAS marker (rs6007600). These findings are consistent with previous reports demonstrating that ancestry-matched LD mapping improves fine-mapping of disease-associated loci and facilitates identification of variants that may be overlooked when European reference panels are used (Wojcik et al. 2019). Our results therefore reinforce the importance of incorporating diverse populations into functional genomic studies to improve the biological interpretation of inherited cancer susceptibility.

Several limitations should be considered when interpreting these findings. First, the functional effects of the prioritized variants were inferred using computational prediction tools and therefore require experimental validation. Luciferase reporter assays, CRISPR-mediated allele editing, RT-qPCR, and protein expression analyses in colorectal cancer cell lines and patient-derived organoids will be important next steps to confirm the predicted allele-specific effects on miRNA-mediated regulation. Second, although the *All of Us* Research Program enabled confirmation of candidate variants in a Hispanic cohort, ancestry-matched transcriptomic, epigenomic, and eQTL datasets for Hispanic CRC remain largely unavailable. Consequently, gene expression, methylation, and survival analyses relied on TCGA-derived resources and publicly available epigenomic datasets, which provide strong biological support but cannot fully capture ancestry-specific regulatory mechanisms (Ardlie et al. 2015). Expanding multiomic resources for Hispanic colorectal cancer represents an important priority for future studies and will substantially improve functional interpretation of inherited risk variants (Bick et al. 2024). Overall, this integrative pipeline establishes an ancestry-anchored framework for decoding non-coding genetic risk, nominating *DCBLD2, GREM1*, and *SMC1B* as priority candidates for functional validation and offering a generalizable approach for investigating post-transcriptional cancer disparities across diverse populations.

## Supplementary Materials

The following are available with this article. Table S1: CRC-associated 3′ UTR GWAS SNPs (SNP loci; positions, risk-allele frequency, p-value, OR/β, trait, PMID). Table S2: CRC-associated SNPs from 453 Hispanic/Latino patient samples. Table S3: AMR LD proxy sets (LDlink; r^2^, D′). Table S4: Ensembl-annotated 3′ UTR variants (SNP id, chromosome position, strand, biotype, gene role in CRC, PMID). Table S5: allele-specific SNP–miRNA binding details (site type, UTR coordinates, context++ score, seed-pairing alignments) for all screened genes. Table S6: full characterization of the remaining candidate genes (TTC22, VIPR1, SMAD9, NT5E, ITPR2, C2orf72), including binding scores.

## Statements and Declarations

### Funding

The research reported here was supported by: (1) Grant # RP210153 from the Cancer Prevention & Research Institute of Texas (CPRIT). (2) Grant # U54MD007592 from the National Institute On Minority Health And Health Disparities of the National Institutes of Health. The content is solely the responsibility of the authors and does not necessarily represent the official views of the National Institutes of Health. (3) Grant # RP230446 from the Cancer Prevention & Research Institute of Texas (CPRIT) to support Md Zahirul Islam Khan.

### Competing Interests

There was no commercial or financial relationship involved in this research. The authors declared no potential conflict of interest.

### Author Contributions

Conceptualization: SSA and MZIK; Methodology: SSA, MZIK and SR; Formal analysis and investigation: SSA and SR; Writing - original draft preparation: SSA and MZIK; Writing - review and editing: SSA, MZIK and SR; Funding acquisition and Supervision: SR. All authors read and approved the final manuscript.

### Data Availability

GWAS Catalog summary statistics are publicly available via the NHGRI-EBI repository. De-identified genomic and electronic health record data from the All of Us Research Program (Controlled Tier Dataset v8) are accessible to authorized researchers through the All of Us Researcher Workbench (https://workbench.researchallofus.org/) under an approved Data User Agreement. All aggregated summary metrics comply with the All of Us Data and Statistics Dissemination Policy. We provide derived data in the Supplementary Materials.

### Ethics approval

This study involved secondary analysis of de-identified genomic and health data from the All of Us Research Program (Controlled Tier Dataset v8) overseen by the NIH All of Us Institutional Review Board (IRB)-UTEP reference number: 1974185. Individual participant e-consent was obtained at enrollment by the All of Us Research Program.

### Consent to participate

Individual participant consent was obtained by the All of Us Research Program at enrollment.

### Consent to publish

Not applicable (no individual patient details, images, or identifying information are included in this manuscript).

## Notes

### Competing Interest Statement

The authors have declared no competing interest.

### Author Declarations

The Institutional Review Board of the University of Texas at El Paso and the Institutional Review Board of the National Institutes of Health All of Us Research Program gave ethical approval for this work (UTEP IRB reference number: 1974185).

